# Effectiveness of immersive VR-based rehabilitation on upper extremity recovery in subacute stroke: a randomized controlled trial

**DOI:** 10.1101/2022.11.08.22281543

**Authors:** Qianqian Huang, Xixi Jiang, Yun Jin, Bo Wu, Andrew D. Vigotsky, Linyu Fan, Pengpeng Gu, Wenzhan Tu, Lejian Huang, Songhe Jiang

## Abstract

**Background and Purpose:** Few effective treatments improve upper extremity (UE) function after stroke. Immersive Virtual Reality (imVR) is a novel and promising strategy for stroke UE recovery. However, research on immersive VR-based UE rehabilitation has been minimal. Here we present a randomized controlled trial to assess the effectiveness of imVR-based UE rehabilitation in patients with subacute stroke and explore the underlying brain network related to imVR-based rehabilitation.

**Methods:** A single-blinded, randomized controlled trial was performed with 40 subjects randomly assigned to either the imVR or the control group (1:1 allocation), each receiving rehabilitation 5 times per week for 3 weeks. Subjects in the imVR group received both immersive VR and conventional rehabilitation, while those in the control group received conventional rehabilitation only. The Fugl-Meyer assessment’s upper extremity subscale (FMA-UE) and the Barthel Index (BI) were our primary and secondary outcomes, respectively. Subjects received assessments and MRI scans at each of the following three time points: immediately after randomization (baseline, Week 0), immediately following conclusion of the randomized rehabilitation program (post-intervention, Week 3), and follow-up 12 weeks after completing the rehabilitation program (follow-up, Week 15). Brain functional connectivity (FC) and a parameter derived from it, degree, were used to assess the performance of immersive VR-based rehabilitation and to relate the change of brain activity to motor recovery.

**Results:** Both intention-to-treat (ITT) and per-protocol (PP) analyses demonstrated the effectiveness of imVR-based UE rehabilitation on subacute stroke. The FMA-UE score was significantly greater in the imVR group compared with the control group at the post-intervention (mean difference: 9.11, 95% CI (1.57-16.64); *p* = 0.019 (ITT); 12.46, 95% CI (4.56 -20.36); *p* = 0.003 (PP)), and at the follow-up (mean difference:11.47, *p* = 0.020 (ITT); 18.85, 95% CI (6.01-31.69); *p* = 0.006 (PP)). The results were consistent for BI scores at the post-intervention (mean difference: 8.28, 95% CI (0.082-16.48); *p* = 0.048 (ITT); 9.77, 95% CI (0.58-18.95); *p* = 0.038 (PP)), and at the follow-up (mean difference: 4.81, 95% CI (0.85-8.77); *p* = 0.019 (ITT); 6.69, 95% CI (0.54-12.84); *p* = 0.034 for (PP)). Moreover, brain functional connectivity analysis found that the motor function improvements are significantly associated with a change in brain functional connectivity in ipsilesional premotor cortex and ipsilesional dorsolateral prefrontal cortex immediately following the intervention and in ipsilesional visual region and ipsilesional middle frontal gyrus after the 12-week follow-up. In addition, a significant increase in the motor recovery rate of the imVR group was observed between the baseline and post-intervention time points (*p* = 0.002).

**Conclusions:** The imVR-based rehabilitation is an effective rehabilitation tool that can improve the recovery of UE functional capabilities of subacute stroke patients when added to standard care. These improvements are associated with distinctive brain reorganizations at two post-stroke timepoints. The study results will benefit future patients with stroke and may provide a new and better method of stroke rehabilitation.

## Introduction

Upper extremity (UE) motor impairment, including loss of movement, sensation, and dexterity, is a common manifestation of patients after stroke [1, 2], compromising patients’ independence in daily activities, thus remarkably diminishing their quality of life. There have been few effective treatments to improve UE function after stroke [3-5]. Moreover, conventional rehabilitation therapy techniques, including motor relearning, proprioceptive neuromuscular facilitation, and neurodevelopmental therapy [6, 7], are tedious and resource-intensive [8]. Thus, developing novel interventions to improve UE function after stroke is of clinical importance and has become one of the top research priorities of stroke identified by scientists and clinicians [9, 10].

Virtual reality (VR) training, an enjoyable and novel rehabilitation strategy, is a promising treatment in that it can enhance motor recovery by providing high-intensity, highly repetitive, and task-orientated training [11, 12], usually unachievable by conventional rehabilitation therapies. VR ranges from non-immersive to fully immersive, depending on the extent to which the users are isolated from the physical environments when interacting with the virtual environment [13]. Non-immersive VR-based rehabilitation has been widely used in stroke rehabilitation for many years and regarded as beneficial for UE recovery [14-16]. However, some researchers failed to demonstrate statistically significant benefits of non-immersive VR-based rehabilitation compared with conventional rehabilitation [17-19]. Instead, they suggested that an immersive VR-based program (imVR) would be a better option due to its unique features: (1) personalized treatment with a variety of training options that are interesting and enjoyable [20]; (2) alternative relaxing environments closer to reality, allowing patients to relearn motor functions in a safe environment [21], and contents are intuitive and easy to operate, therefore boosting patients’ abilities to transfer the skills learned in the virtual environment into real life [22, 23].

So far, the use of imVR systems for UE motor rehabilitation has not yet been sufficiently studied or implemented. While a few studies have applied imVR in stroke rehabilitation training and demonstrated that it could improve the effectiveness of UE rehabilitation training in stroke patients [24-26], most of these studies have small sample sizes (even single cases) and are without a control group, only focus on short-term effects, and seldom explore the underlying brain mechanisms [25, 27, 28]. Additionally, studies evidence that recovery processes plateau after about 6 months [29]; since neuroplasticity may have become less elastic in this timeframe [30], patients in the subacute phase (7 days – 6 months post-stroke) [31] may benefit from VR therapies more than in chronic phases of stroke [30]. Thus, performing a randomized trial with a control group is necessary to compare the effectiveness of imVR with conventional therapy on UE motor recovery in stroke patients in their subacute phase.

In this study, we hypothesized that imVR rehabilitation for patients with subacute stroke would result in better UE motor recovery than conventional therapy. Subsequently, this improvement would correlate with brain neurophysiological change. To assess this hypothesis, we enrolled 40 patients in a single-blind, parallel-group, and randomized trial, during which fMRI was used to investigate neuroplasticity resulting from rehabilitation [32] and the relationship between brain functional topologies and recovery of UE motor performance was explored afterward, expectedly elucidating underlying brain mechanisms of imVR-based rehabilitation.

## Materials and methods

### Study design

This study aimed to assess the effectiveness of immersive VR-based UE rehabilitation on patients with subacute stroke. As shown in **Fig. 1**, 85 stroke patients in their subacute stage were recruited, and 40 subjects were enrolled in a single-blinded randomized controlled trial. They were randomly evenly allocated into a new rehabilitation treatment with an immersive VR system (imVR group) or a conventional treatment program (Control group). Each subject was randomly assigned a code based on computer-generated, permuted block randomization with a block size of 4. Because of the nature of the intervention, subjects and therapists could not be blinded to the allocated treatment. These therapists did not participate in assessments of the outcomes. The assessments include MRI scans and evaluations performed by assessors who are blinded to this study.

**Figure 1.**
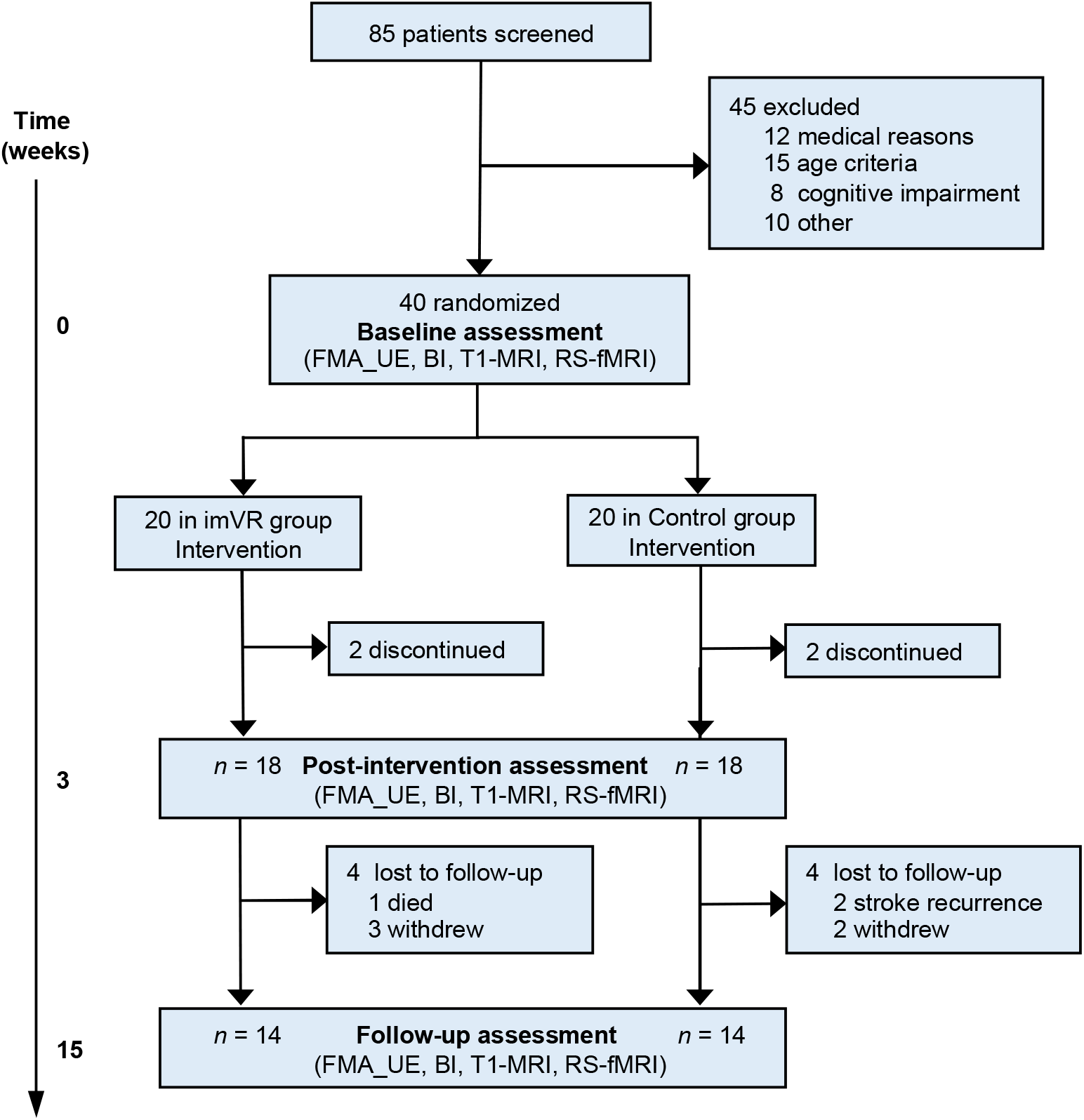
Trial profile. Week 0: baseline assessment; Week 0 ∼ Week 3: intervention in the imVR or the Control groups; Week 3: post-intervention assessment; Week 15: follow-up assessment. imVR: immersive virtual reality; FMA-UE: Fugl-Meyer Assessment-Upper Extremity; BI: Barthel Index

The inclusion criteria were as follows: to be eligible, subjects must (1) be over 30 but less than 85 years old; (2) have had their first stroke within the past month; (3) be at the subacute stage with a subcortical lesion location including the basal ganglia, internal capsule, corona radiata or brainstem; and (4) have a starting upper-limb function of Brunnstrom [37] stage II–IV. The exclusion criteria were as follows: (1) history of transient ischemic attack (TIA); (2) failure of critical organs, such as heart, lung, liver, and kidney; (3) previous history of brain neurosurgery or epilepsy; (4) severe cognitive impairments or aphasia (incapable of understanding the instructions given by therapists); (5) not suitable for an MRI scan; and (6) enrollment in another clinical trial involving physical therapy or an investigational drug.

Subjects received assessments at the following three time points: immediately after randomization (baseline, Week 0), immediately following conclusion of the randomized rehabilitation program (post-intervention, Week 3) and follow-up 12 weeks after conclusion of the rehabilitation program (follow-up, Week 15).

This study was performed at the Second Affiliated Hospital and Yuying Children’s Hospital of Wenzhou Medical University, China from March 2017 to July 2021. The study was approved by the Institutional Review Board of the Hospital (No. 2017LCKY-09) and all participants provided written informed consent. The clinical trial was registered at ClinicalTrials.gov (NCT03086889) and its original study protocol has been published in [33].

### Procedures

Subjects in the Control group received a 60-minute conventional rehabilitation program per day. This conventional rehabilitation program consists of physical and occupational therapy, including grips and selective finger movements, gross movement, strength training, stretching, and training in activities of daily life. In contrast, subjects in the imVR group received the first 30 minutes of conventional rehabilitation, and in the second 30 minutes, the rehabilitation was performed in imVR systems. All subjects received rehabilitation training 5 days per week over 3 weeks.

We used the HTC Vive-VR HMD system as the immersive VR device. Subjects in the imVR group were required to complete 6 programs: frying dumplings and noodles by controlling a wok handle in a virtual kitchen (**Supplementary Fig. 1A**); popping balloons by controlling a sword in a virtual fencing hall (**Supplementary Fig. 1B**); punching dolls by controlling a big fist in a virtual boxing arena (**Supplementary Fig. 1C**); playing basketball in a virtual court, in which the ball is shot by a controller and the height and distance is varied over time (**Supplementary Fig. 1D**); collecting eggs into a virtual basket by a controller (**Supplementary Fig. 1E**); and tidying up a desk and moving objects to a designated position in a virtual office (**Supplementary Fig. 1F**).

In the early stages of rehabilitation, due to the poor function of the hemiplegic side upper limb, subjects had to complete the imVR programs with the help of the limb on the unaffected side. With the recovery of the hemiplegic side upper limb function, the subjects independently completed the 6 games with only their limb on the hemiplegic side.

### Outcomes

The upper extremity portion of the Fugl-Meyer assessment (FMA-UE) [34] was the primary outcome measure for this trial. The FMA-UE, which measures arm movement ability across several domains (motor function, balance, sensation, range of motion, and pain), is a standard clinical tool for evaluating changes in motor impairment after stroke [34]. The Barthel Index (BI) [35], the secondary outcome measure, measures activities of daily living (ADL), consisting of feeding, grooming, bathing, bowel control, chair transfer, bladder control, toileting, dressing, ambulation and stair climbing. RS-fMRI was an additional outcome measure. Brain functional connectivity (FC) and its derivative parameter, degree, were used to assess neuro-biological correlates of VR-based rehabilitation and to relate changes in brain activity to motor recovery.

### MRI data acquisition

All subjects were scanned on a 3.0 T GE-Discovery 750 scanner with the following parameters: for anatomical T1-MRI data: TR/TE = 7.7/3.4 ms, flip angle = 12°, FOV = 256 mm × 256 mm, resolution = 256×256, number of slices = 176, isometric voxel size = 1 × 1 × 1 mm; for fMRI data: TE/TR = 30/2500 ms with interleaved ordering, voxel size = 3.4375 × 3.4375 × 3.5 mm^3^, in-plane resolution = 64 × 64, number of volumes = 230, and flip angle = 90°.

### RS-fMRI data quality control, preprocessing and registration

Mean framewise displacement (mFD) of each RS-fMRI data set, calculated as the sum of mean displacement along 6 dimensions, indicating the extent of head motion over the duration of the scan, was used as a metric of quality control of RS-fMRI data and a confound in the further statistical analysis.

A similar preprocessing pipeline to [36] was applied to all RS-fMRI data and briefly introduced as follows: removal of the first 4 volumes (10 seconds) for magnetic field stabilization; motion correction; slice-time correction; intensity normalization; high-pass temporal filtering (0.008 Hz) for correcting low-frequency signal drift; nuisance regression of 6 motion vectors, signal-averaged overall voxels of the eroded white matter and ventricle region, and global signal of the whole brain; motion-volume censoring by detecting volumes with a FD larger than 0.5 millimeters, Derivative Variance Root mean Square after Z normalization larger than 2.3, and standard deviation after Z normalization larger than 2.3, and scrubbing above detected (volume = *i*) and adjacent four volumes (*i* − 2, *i* − 1, *i* + 1, *i* + 2) [37, 38]; band-pass filtering (0.008-0.1 Hz) by applying a 4^th^-order Butterworth filter.

All pre-processed RS-fMRI data were registered to the MNI152 template using a two-step procedure, in which the mean of preprocessed fMRI data was registered with a 7-degree-of-freedom affine transformation to its corresponding T1 brain (FLIRT); transformation parameters were computed by nonlinearly registering individual T1 brain to the MNI152 template (FNIRT). Combining the two transformations by multiplying the matrices yielded transformation parameters normalizing the pre-processed fMRI data to the standard space. All the final registered images were manually examined.

After the registration, for those subjects who had left-sided lesions, the registered images were flipped from left to right along the midsagittal line. In the end, the right side corresponded to the ipsilesional hemisphere.

### Resting-state functional connectivity networks

For each subject, their resting-state brain functional connectivity networks (FCN) covering gray matter were generated. First, blood oxygenation level dependent (BOLD) signal was extracted from each gray matter voxel in the registered RS-fMRI data. Following this, we calculated voxel-based pairwise Pearson correlation coefficients of BOLD signals to construct a correlation matrix, which was then Fisher’s z transformed. To normalize the variation of strength of brain FCN across individuals, a link density — the percentage of links with respect to the maximum number of possible links — was predetermined, corresponding to a threshold [39, 40]. In our study, 10% link density was applied. Consequently, an indirectly connected brain FCN was generated after the correlation matrix was binarized by the subject-dependent threshold to create an adjacency matrix.

### Degree comparison and association between changes in degree and motor recovery

We used degree to investigate the effect of imVR rehabilitation training on brain activity. For each subject, their degree map was generated from the brain FCN by calculating the number of functional links of each gray matter voxel. The links within two adjacent voxels were excluded due to the effect of motion [38, 41]. To compare degrees between the imVR and the Control groups, we used a general linear model (GLM) with degree at post-intervention and at follow-up, respectively, as the dependent variable, the two groups as the independent variable, baseline degree, age, sex, side of brain lesion, time since stroke, hypertension, diabetes, and mFD as confounds; family-wise cluster correction (*t* value > 3.5, *p* < 0.01) [42] was applied afterward.

For each statistically significant cluster, we compared the average degree count extracted from the cluster between the imVR and the Control groups and healthy controls (HCs), respectively, to investigate the difference relative to HCs. As shown in **Supplementary Table 1**, these HCs are from the same data set as in [43] — they were matched on age, sex, and brain motion during scanning, represented by *log(*mFD). In addition, the association analyses between change in degree extracted from the cluster and change of outcomes were performed.

### Network reorganization

After identifying brain clusters that statistically significantly differed between groups, we explored which regions these significant clusters connected to and if these connected regions were also statistically significantly different (reorganized) between the imVR and the Control groups. The analysis was run in network (module) space spanned by 333 cortical parcels defined in [44] and 16 in-house-defined subcortical regions, from which 13 functional networks are constructed: visual, auditory, default-mode, cingulo-opercular task control, fronto-parietal task control, sensory/somatomotor mouth, sensory/somatomotor hand, dorsal attention, ventral attention, subcortex, salience, cinguloparietal, retrosplenial temporal [44]. For each subject, the status of functional connectivity between each significant cluster and the 349 parcels (regions) was set to as “*connected*” if their correlations coefficients were greater than the subject-dependent threshold corresponding to 10% link density or “*disconnected*” if less, generating a vector of 349 connection status. Chi-squared test was independently applied to each parcel to determine if there existed statistically significant difference of connection status between the imVR and the Control groups (*p* value < 0.05). The results were reported in circular plots.

### Statistical and data analyses

In our previous sample size calculation [33], we estimated that 30 subjects per group would be sufficient to assess the effectiveness of the imVR training given a two-tailed comparison and set the type I error rate at 0.05 with 80% power and effect size of 0.75. In our protocol, one interim analysis was planned after 60% of subjects completed the post-intervention. If the *P* value corresponding to the effectiveness of the imVR (FMA-UE) was less than 0.025, the trial would be terminated earlier.

Both intention-to-treat (ITT) and per-protocol (PP) analyses were performed to assess the effectiveness of the trial. The ITT analysis was conducted with all randomly assigned participants included in the analysis, applying the Markov Chain Monte Carlo method with linear regression (only FMA-UEs or BIs as predictors in the model) for the imputation of any missing value (20 subjects in imVR vs. 20 subjects in Control). The PP analysis included participants who had at least a 2-week long intervention (18 vs. 18 post-intervention and 14 vs. 14 follow-up).

This randomized controlled trial is a two-group independent design examining the effects of imVR on rehabilitation of patients with subacute stroke and the assessments were repeated three times. We were interested in the change of outcomes (i.e., recovery) between the two groups. So, for both the FMA-UE /the BI, a one-way analysis of covariance (ANCOVA) model was used, with the FMA-UE /BI at post-intervention or at follow-up, respectively, as the dependent variable, the two groups as the independent variable, baseline FMA-UE score/BI, age, sex, site, time since onset, hypertension and diabetes as covariates. *P* < 0.05 was statistically significant.

Independent *t*-tests and Mann Whitney U tests were employed to compare outcomes that did and did not meet residual normality assumptions, respectively, between the two groups. The Chi-square test was used to compare categorical outcomes.

A mixed model was used to assess the significance of difference of brain motion during scanning, represented by *log*(mFD), across three assessments between the imVR and the Control groups. *P* < 0.05 was statistically significant.

To investigate the effects of imVR on the brain both at post-intervention and the follow-up, for each time point the GLM model was applied with degree as the dependent variable, two groups as the independent variable, base-line degree, age, sex, site, time since onset, hypertension, diabetes, and *log*(mFD) as covariates; cluster-correction was performed afterwards (*t* value > 3.5, *p* < 0.01).

Pearson correlations were performed to examine how degree, extracted from the significant cluster, tracks with changes of outcomes (FMA-UE or BI) using (1) post-intervention and baseline and (2) follow-up and baseline, respectively. *P* < 0.05 was statistically significant.

All statistical analyses were performed using MATLAB 2016a and SPSS 23 (IBM Corp. in Armonk, NY).

## Results

### Baseline Characteristics of Participants

As shown in **Figure. 1**, after 45 patients were excluded, a total of 40 participants were randomized in this study and evenly assigned to the imVR group (*n* = 20) or the Control group (*n* = 20). 2 participants in each group withdrew from the study and 4 per group were lost to follow-up between the 3-week intervention and follow-up assessment. The largest area of lesion at the axial cross section of participants for brain image analysis is marked by a yellow arrow in **Supplementary Fig. 2**. Demographic and baseline clinical characteristics of both groups were summarized in **Table 2**. For the RS-fMRI image quality, mFD of all subjects including HCs was less than 0.2 mm [41]; as shown in **Supplementary Fig. 3**, a mixed model determined that there was no statistically significant difference in brain motion during scanning, represented by *log*(mFD), across the three assessments between the imVR and the Control groups (*p* = 0.806).

**Table 1.** Demographic and baseline clinical characteristics of enrolled 40 subjects.

|  | imVR (*n* = 20) | Control (*n* = 20) |
| --- | --- | --- |
| **Age** (*years*) mean (*SD*) | 63.30 (14.32) | 65.05 (6.14) |
| **Sex** (male) *n* (%) | 13 (65.00) | 11 (55.00) |
| **Right handedness** *n* (%) | 20 (100) | 20 (100) |
| **Time since stroke** (*days*) mean (*SD*) | 18.80 (8.44) | 19.00 (6.64) |
| **Hypertension** *n* (%) | 13 (65.00) | 16 (80.00) |
| **Diabetes** *n* (%) | 11 (55.00) | 12 (60.00) |
| **Stroke type** *n* (%) |  |  |
| ischemia | 19 (95.00) | 18 (90.00) |
| hemorrhage | 1 (5.00) | 2 (10.00) |
| **Side of brain lesion** *n* (%) |  |  |
| right | 10 (50.00) | 13 (65.00) |
| left | 10 (50.00) | 7 (35.00) |
| **FMA-UE** mean (*SD*) | 17.50 (12.84) | 14.75 (12.97) |
| **BI** mean (*SD*) | 56.00 (17.44) | 47.50 (21.00) |

SD: Standard Deviation; imVR: immersive Virtual Reality; FMA-UE: Fugl-Meyer Assessment-Upper Extremity; BI: Barthel Index.

**Table 2.** Outcomes at baseline, post-intervention and follow-up by groups.

|  | imVR | Control | Between group comparisons |  |
| --- | --- | --- | --- | --- |
|  | Outcome | Outcome | Mean difference (95% CI) | *p* value |
| **ITT analysis** |  |  |  |  |
| **FMA-UE** mean (*SD*) |  |  |  |  |
| Baseline | 17.50 (12.84) | 14.75 (12.97) | 2.75 (-5.51 to 11.01) |  |
| Post-intervention | 34.90 (18.44) | 23.95 (20.67) | 9.11 (1.57 to 16.64)<sup>b</sup> | **0.019<sup>c</sup>** |
| Follow-up | 47.95 (15.11) | 36.00 (19.96) | 11.47 (1.93 to 21.02)<sup>b</sup> | **0.020<sup>c</sup>** |
| **BI** mean (*SD*) |  |  |  |  |
| Baseline | 56.00 (17.44) | 47.50 (21.00) | 8.50 (-3.86 to 20.86) |  |
| Post-intervention | 85.25 (12.19) | 72.75 (21.67) | 8.28 (0.082 to 16.48)<sup>b</sup> | **0.048<sup>c</sup>** |
| Follow-up | 98.75 (3.40) | 92.20 (8.22) | 4.81 (0.85 to 8.77)<sup>b</sup> | **0.019<sup>c</sup>** |
| **PP analysis** |  |  |  |  |
| **FMA-UE** mean (*SD*) |  |  |  |  |
| Baseline | 16.00 (12.09) | 14.17 (12.70) | 1.83 (-6.57 to 10.23) | 0.660<sup>a</sup> |
| Post-intervention | 33.67 (18.16) | 22.39 (19.60) | 12.46 (4.56 to 20.36)<sup>b</sup> | **0.003<sup>c</sup>** |
| Follow-up | 48.57 (14.98) | 35.21 (21.52) | 18.85 (6.01 to 31.69)<sup>b</sup> | **0.006<sup>c</sup>** |
| **BI** mean (*SD*) |  |  |  |  |
| Baseline | 54.17 (16.91) | 48.61 (21.88) | 5.56 (-7.69 to 18.80) | 0.400<sup>a</sup> |
| Post-intervention | 84.17 (12.28) | 72.22 (22.83) | 9.77 (0.58 to 18.95)<sup>b</sup> | **0.038<sup>c</sup>** |
| Follow-up | 99.64 (1.34) | 91.79 (9.53) | 6.69 (0.54 to 12.84)<sup>b</sup> | **0.034<sup>c</sup>** |

Data are shown for the ITT analysis (20 subjects in imVR vs. 20 subjects in Control) and for the PP analysis (18 vs. 18 post-intervention and 14 vs. 14 follow-up). FMA-UE: Fugl-Meyer Assessment-Upper Extremity; BI: Barthel Index; CI: Confidence Interval; imVR: immersive Virtual Reality; ITT: intention to treat; PP: per-protocol. a: *p* value was determined using independent samples *t*-test. b: adjusted estimates after controlling for baseline FMA-UE score or BI score, age, gender, site, time since onset, hypertension and diabetes. c: *p* value was determined using ANCOVA model with baseline score, age, sex, side of brain lesion, time since stroke, hypertension and diabetes as covariates of no interest.

### Motor performance

The ITT analysis demonstrated that the primary outcome, FMA-UE score, was statistically significantly greater in the imVR group compared with the Control group both at the post-intervention (adjusted effect: 9.11, 95% CI (1.57 – 16.64); *p* = 0.019) and at the follow-up assessment (adjusted effect: 11.47, 95% CI (1.93 – 21.02); *p* = 0.020) (**Table 2**, top). The results are consistent with the PP analysis (**Table 2**, bottom) at the post-intervention (adjusted effect: 12.46, 95% CI (4.56 – 20.36); *p* = 0.003) and at the follow-up assessment (adjusted effect: 18.85, 95% CI (6.01 – 31.69); *p* = 0.006).

Similarly, both analyses showed that the secondary outcome, BI score, was also statistically significantly greater in the imVR group compared with the Control group both at the post-intervention (adjusted effects: 8.28, 95% CI (0.082 – 16.48); *p* = 0.048 for ITT analysis and 9.77, 95% CI (0.58 - 18.95); *p* = 0.038 for PP analysis) and at the follow-up (adjusted effects: 4.81, 95% CI (0.85 – 8.77); *p* = 0.019 for ITT analysis and 6.69, 95% CI (0.54 – 12.84); *p* = 0.034 for PP analysis).

**Fig. 2A** depicts the individual FMA-UE scores at baseline, post-intervention, and follow-up, illustrating (1) from baseline to post-intervention, the FMA-UE scores increased across both groups, but the slopes of the imVR group (red lines) were generally steeper than the Control group (blue lines); (2) from post-intervention to follow-up, FMA-UE scores continued to increase, but the slopes were not as steep as those in previous stage for both groups. These observations were statistically supported by the results of **Fig. 2B**, where bar plots of the group means (± standard error) shows a significant increase of FMA-UE change rate for the imVR group after the training (*p* = 0.002) but no significant difference between post-intervention and follow-up (*p* = 0.303).

**Figure 2.**
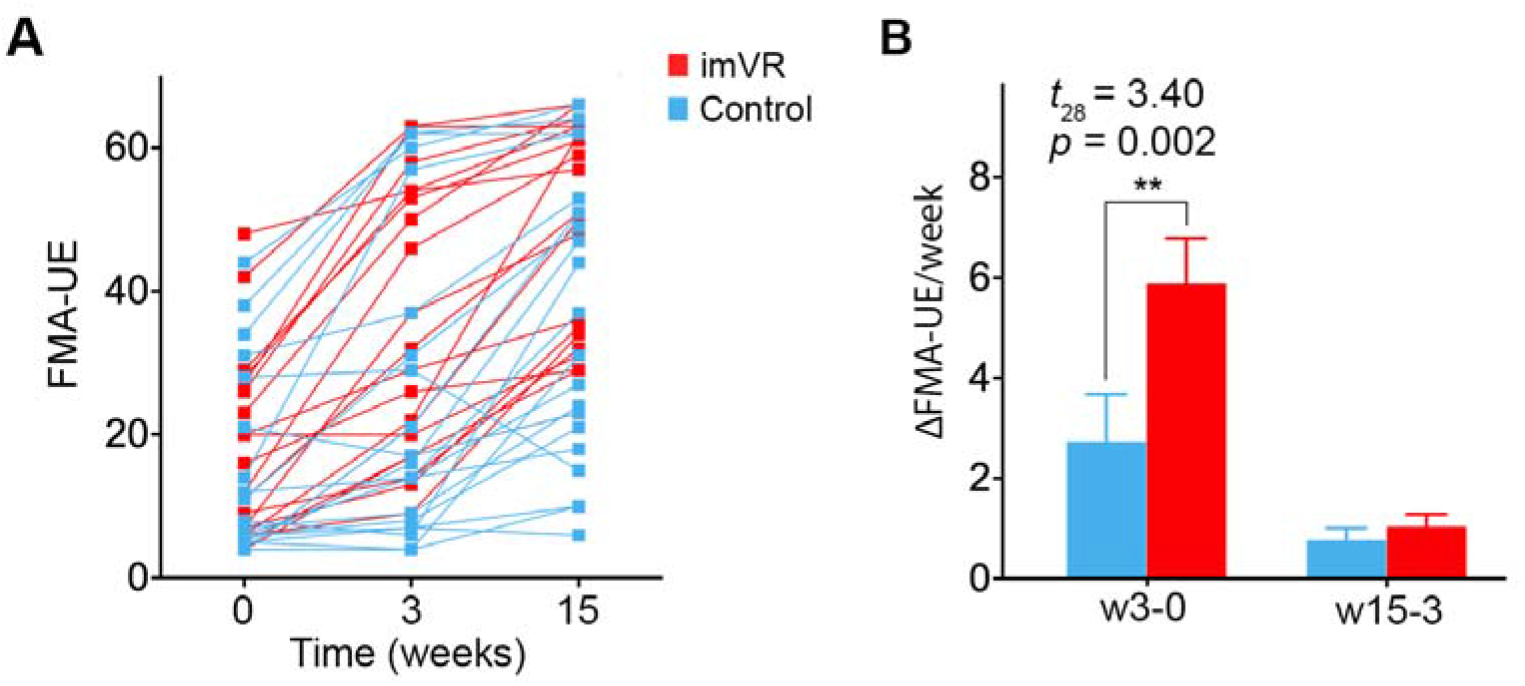
Comparison of motor recovery rate between training and follow-up. (**A**) Dot plot depicts individual FMA-UE score at the baseline (Week 0), post-intervention (Week 3), and follow-up (Week 15). (**B**) Bar plot (mean and SE) shows a significant increase of FMA-UE change rate for the imVR group after the training (*t*_28_ = 3.40, *p* = 0.002). *p* value was determined using an ANCOVA model with age, sex, side of brain lesion, time since stroke, hypertension and diabetes as covariates of no interest. FMA-UE: Fugl-Meyer Assessment-Upper Extremity; ΔFMA-UE: the change of FMA-UE between two time points; w3-0: duration between Week 3 and Week 0; w15-3: duration between Week 15 and Week 3; imVR: immersive virtual reality.

### Post-intervention fMRI results

After cluster-correction (*t* >□3.5 and *p*□<□0.01), compared to the Control group, the imVR group exhibited greater degrees in IL_PMd (*p* = 0.008) and IL_M1 (*p* = 0.003) regions (**Table 3, Fig. 3A, Supplementary Fig. 4A**), and lower degrees in IL_and CL_DLPFC (*p* = 0.003; *p* < 0.001) regions at the end of intervention (Week 3) (**Table 3, Fig. 3E, Supplementary Fig. 4B**).

**Figure 3.**
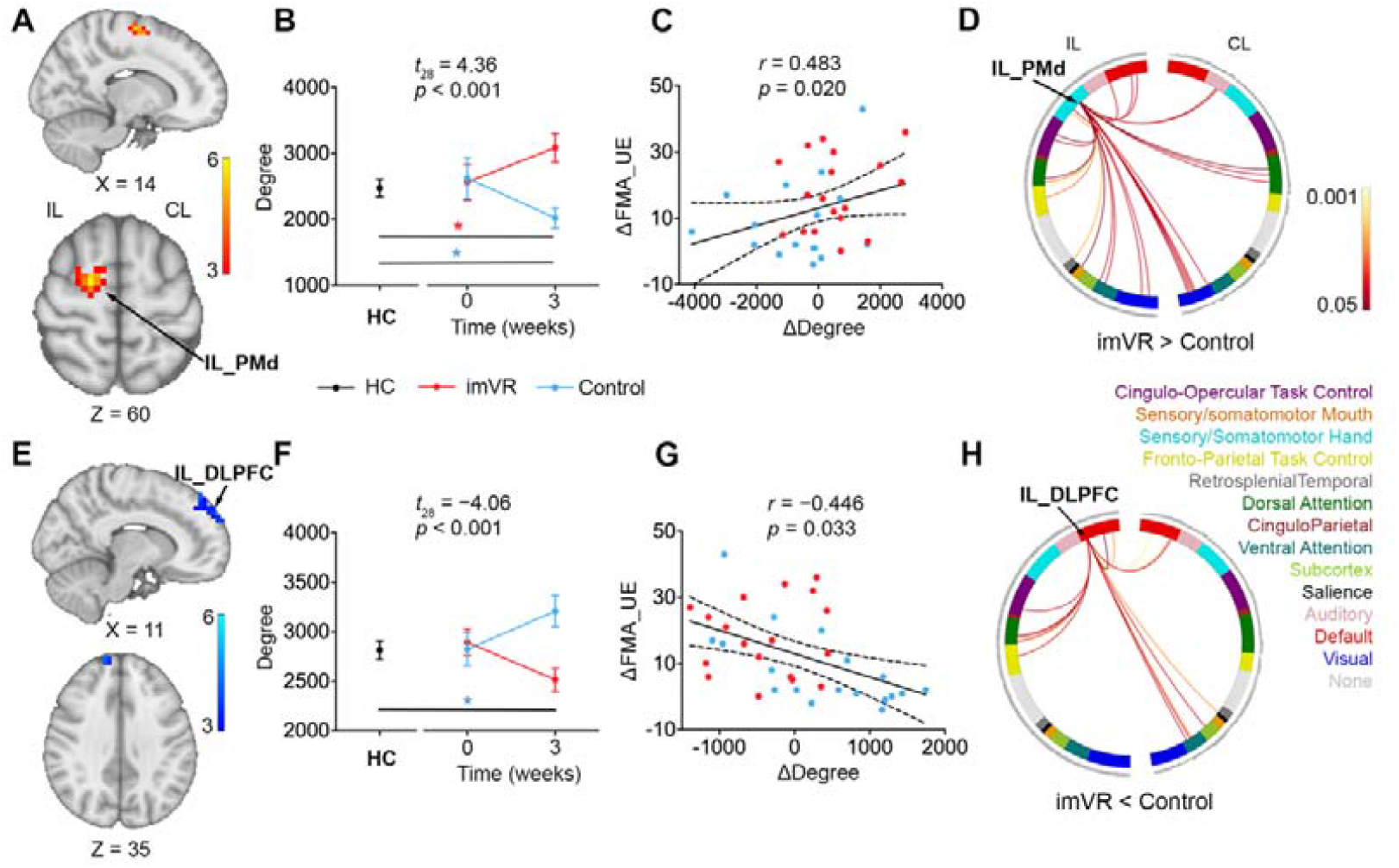
Changes in functional connectivity of the ipsilesional premotor cortex and ipsilesional dorsolateral prefrontal cortex significantly associated with recovery of motor performance after the intervention. **(A)** The imVR group has greater degree in IL_PMd region at the end of intervention compared with the Control group (cluster-corrected, *t* >□3.5, *p*□<□0.01). **(B)** Post hoc analysis indicates the greater IL_PMd degree in the imVR group compared to the Control group (*t*_28_ = 4.36, *p* < 0.001) at the end of the intervention. Compared with HCs, IL_PMd’s was increased (*p* = 0.014) and decreased (*p* = 0.035) from the baseline to the post-intervention for the imVR and the Control groups, respectively. Data plotted as mean ± SE. **(C)** A positive correlation between the change in IL_PMd’s degree and the change of FMA-UE (*r* = 0.483, *p* = 0.020) from baseline to post-intervention. **(D)** Circular plot shows the difference of functional connections to IL_PMd between the imVR and Control groups (*p* < 0.05) in the network space. Most connection differences are from the sensory/somatomotor hand, visual, fronto-parietal task control, ventral attention, dorsal attention, default and cingulo-opercular task control networks on the ipsilesional hemisphere. Line color is proportional to statistical incompatibility with the null (0.001 < *p* < 0.05). The 14 networks and colors associated with the networks are defined in [45] and are illustrated below the circle. **(E)** The imVR group presents had lower IL_ DLPFC degree at the end of intervention compared with the Control group (Week 3) (cluster-corrected, *t* >□3.5, *p*□<□0.01). **(F)** Post hoc analysis greater mean degree in IL_DLPFC in Controls than the imVR group (*t*_28_ = 4.06, *p* < 0.001) at the end of the intervention. Compared with HCs, the Control group had greater mean degree in IL_DLPFC (*p* = 0.028). Data plotted as mean ± SE. **(G)** Changes in mean degree of IL_DLPFC correlated with changes of FMA-UE (*r* = -0.446, *p* = 0.033) from baseline to post-intervention. **(H)** Circular plots show the difference of functional connections to IL_DLPFC between the Control and imVR groups (*p* < 0.05) in the network space. Most of the differences in connections are from the ventral attention and DMN on contralesional hemisphere, and DMN, cingulo-opercular task control, frontal-parietal task control and dorsal attention network on ipsilesional hemisphere. ΔDegree = post-intervention degree minus baseline degree; ΔFMA-UE=post-intervention FMA-UE minus baseline FMA-UE; IL: Ipsilesional; CL: Contralesional

**Table 3.** Brain regions with statistically significant local degree differences between baseline and post-intervention at 10% link density

| Brain regions<sup>a</sup> | Side | Peak MNI coordinates |  |  | Cluster size (mm<sup>3</sup>) | *p* value<sup>b</sup> |
| --- | --- | --- | --- | --- | --- | --- |
|  |  | X | Y | Z |  |  |
| imVR > Control |  |  |  |  |  |  |
| PMd | IL | 18 | -6 | 60 | 2304 | **0.008** |
| M1 | IL | 34 | -26 | 60 | 2688 | **0.003** |
| imVR < Control |  |  |  |  |  |  |
| DLPFC | IL | 14 | 58 | 32 | 2816 | **0.003** |
| DLPFC | CL | -14 | 58 | 36 | 3584 | **< 0.001** |

**Abbreviations:** PMd: Dorsal Premotor Cortex; M1: Primary Motor Cortex; DLPFC: Dorsolateral Prefrontal Cortex; IL: Ipsilesional; CL: Contralesional; MNI: Montreal Neurological Institute; imVR: immersive virtual reality.
**Note:** <sup>a</sup>Harvard-Oxford cortical and subcortical structural atlas; <sup>b</sup>cluster-corrected ( $t > 3.5$ , $p < 0.01$ ).

As shown in **Fig. 3B** and **Fig. 3F**, post hoc analyses indicated that the imVR group presents significantly higher mean degree in IL_PMd (*t*_28_ = 4.36, *p* < 0.001) and lower mean degree in IL_DLPFC (*t*_28_ = -4.06, *p* < 0.001) than the Control group at the post-intervention; compared with HCs, the mean degree in IL_PMd was increased (*p* = 0.014) and decreased (*p* = 0.035) for the imVR and the Control groups, respectively, while IL_DLPFC was increased (*p* = 0.028) for the Control group only. As shown in **Fig. 3C** and **Fig. 3G**, the post hoc analyses also revealed that the change of mean degree in IL_PMd and IL_DLPFC between the post-intervention and the baseline is, respectively, positively and negatively correlated with changes in FMA-UE (*r* = 0.483, *p* = 0.020; *r* = −0.446, *p* = 0.033), indicating that the change of functional connectivity in both IL_PMd and in IL_DLPFC is associated with recovery of motor performance after the intervention. Furthermore, in network space, as shown in **Fig. 3D**, for IL_PMd, which is assigned to the sensory/somatomotor hand network, most of the degree differences (more connections to IL_PMd in the imVR group than in the Control group) are from the sensory/somatomotor hand, visual, frontal-parietal task control, ventral attention, dorsal attention, default mode network (DMN) and cingulo-opercular task control networks on the ipsilesional hemisphere. For IL_DLPFC, as shown in **Fig. 3H**, which is assigned to DMN, most of the degree difference (more connections to IL_DLPFC in the Control group than in the imVR group) are from the ventral attention and DMN on contralesional hemisphere, and DMN, cingulo-opercular task control, frontal-parietal task control and dorsal attention network on ipsilesional hemisphere.

The results of the other two statistically significant regions found at the end of intervention, IL_M1 and CL_DLPEC, are shown in **Supplementary Fig. 4**, the details of which can be found in the supplementary post-intervention fMRI results.

### Follow-up fMRI results

At the end of the follow-up (Week 15), after cluster-correction (*t* >□3.5 and *p*□<□0.01), the imVR group exhibited greater degrees in IL_V1 (*p* = 0.002), CL_V1 (*p* < 0.001), CL_SPG (*p* < 0.001) and IL_LOC (*p* < 0.001) (**Table 4, Fig. 4A, Supplementary Fig. 5A**), and the lower degree in IL_MFG (*p* < 0.001), IL_PMv (*p* = 0.004), IL_IFG (*p* < 0.001), CL_mPFC (*p* < 0.001) and CL_FP (*p* < 0.001) regions as compared to the Control group (**Table 4, Fig. 4E, Supplementary Fig. 5B**). These 9 regions do not overlap with any of the 4 regions found to be statistically significant at the post-intervention timepoint.

**Figure 4.**
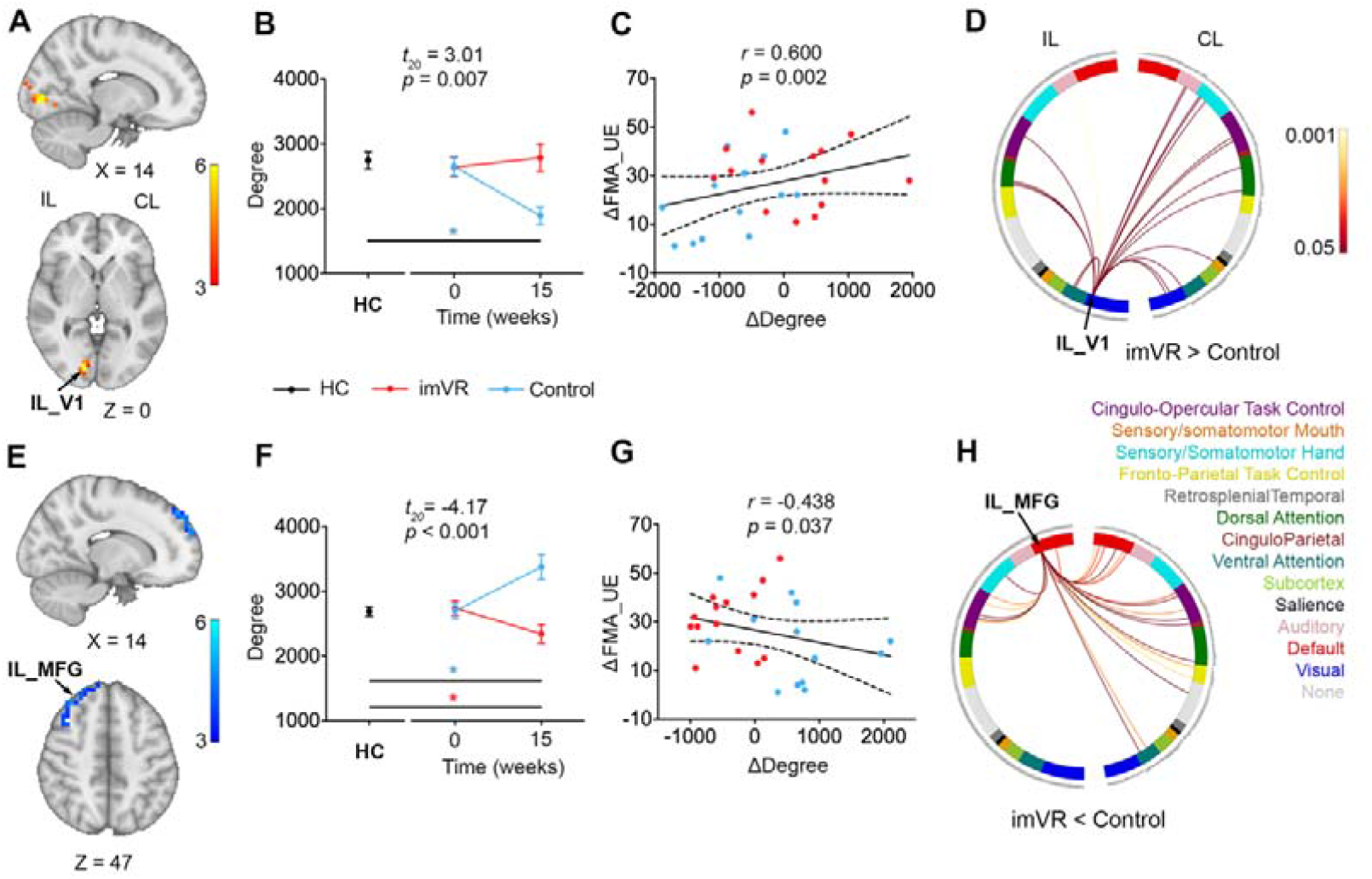
Changes in functional connectivity of the ipsilesional visual region and ipsilesional dorsolateral prefrontal cortex significantly associated with recovery of motor performance at the end of the follow-up. **(A)** imVR had greater IL_V1 degree at the end of the follow-up compared with the Control group (cluster-corrected, *t* >□3.5, *p*□<□0.01). **(B)** imVR had greater IL_V1 mean degree than the Control group (*t*_20_ = 3.01, *p* = 0.007) at the end of the follow-up. Compared with HCs, the mean degree of IL_V1 was decreased (*p* < 0.001) from the baseline to the follow-up for the Control group. Data plotted as mean ± SE. **(C)** Changes in the mean degree of IL_V1 are correlated with changes in FMA-UE (*r* = 0.600, *p* = 0.002) from baseline to the end of the follow-up. **(D)** Functional connections to IL_V1 differed between the imVR and Control groups (*p* < 0.05) in the network space. Most of the differences were from the sensory/somatomotor hand, visual, auditory, cingulo-opercular task control, dorsal attention and ventral attention networks in the contralesional hemisphere. **(E)** imVR had lower degree in IL_MFG region at the end of the follow-up compared with the Control group (cluster-corrected, *t* >□3.5, *p*□<□0.01). **(F)** The Control group had greater mean degree in IL_MFG than the imVR group (*t*_20_ = -4.17, *p* < 0.001) at the end of follow-up. Compared with HCs, the mean degree in IL_MFG was increased (*p* = 0.021) and decreased (*p* < 0.001) for the imVR and the Control groups, respectively. Data plotted as mean ± SE. **(G)** Mean IL_MFG degree was negatively correlated between change of mean degree in IL_MFG and change of FMA-UE (*r* = -0.438, *p* = 0.037) at the end of the follow-up. **(H)** Circular plots show differential functional connections to IL_DLPFC between the Control and imVR groups (*p* < 0.05) in the network space. Most of the differences in connections are from the ventral attention, default, fronto-parietal task control, cingular-opercular task control networks on the contralesional hemisphere. ΔDegree = follow-up degree minus baseline degree; ΔFMA-UE = follow-up FMA-UE minus baseline FMA-UE; IL: Ipsilesional; CL: Contralesional

**Table 4.**
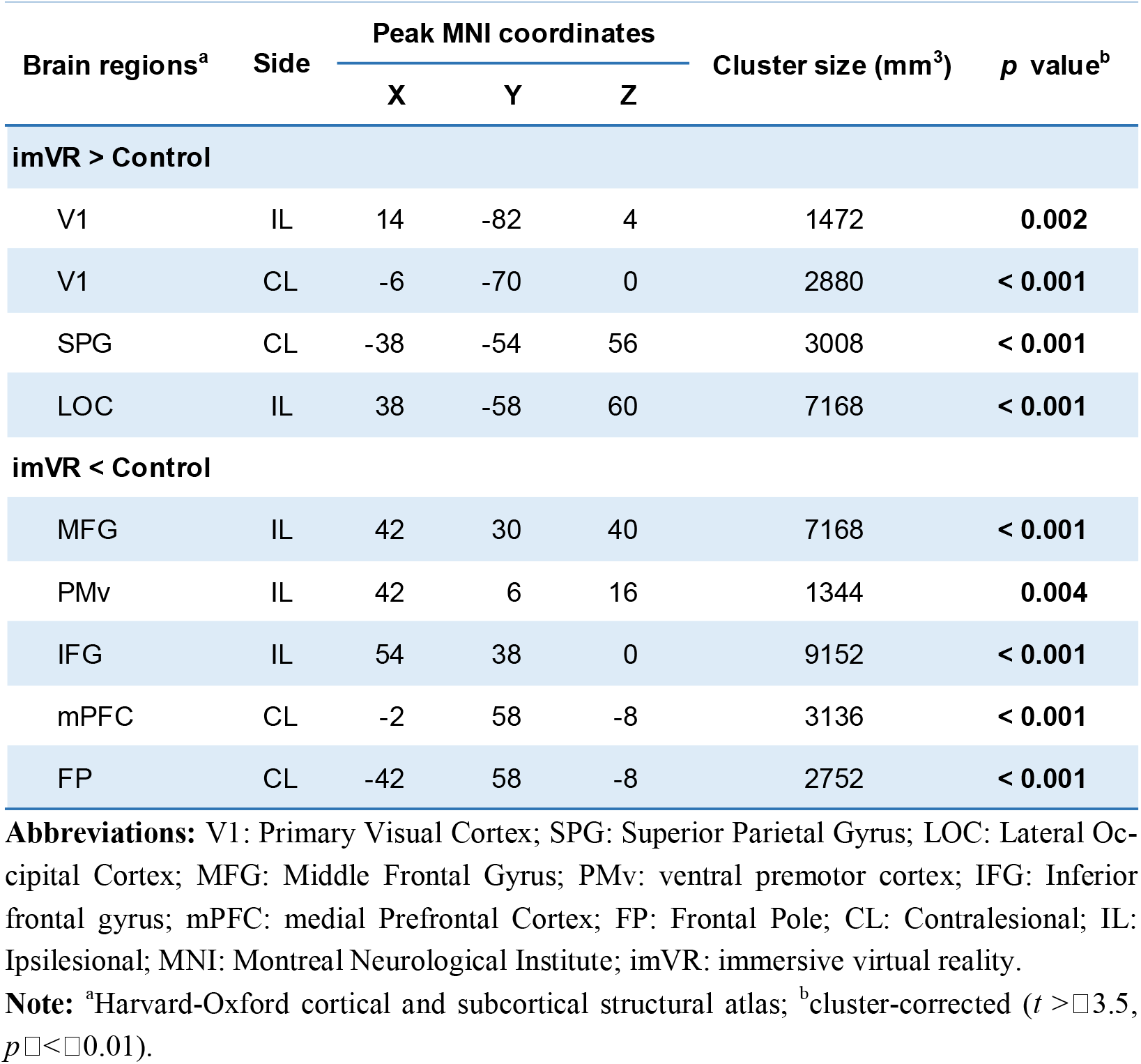
Brain regions with statistically significant local degree differences between baseline and the end of follow-up at 10% link density

Post hoc analyses indicated greater mean degree in IL_V1 (*t*_20_ = 3.01, *p* = 0.007) and lower mean degree in IL_MFG (*t*_20_ = -4.17, *p* < 0.001) in imVR as compared to the Control group at the end of the follow-up; compared with HCs, the mean degree in IL_MFG is was decreased (*p* = 0.021) and increased (*p* < 0.001) from the baseline to the end of follow-up for the imVR and the Control groups, respectively, while that in IL_V1 was lower (*p* < 0.001) in the Control group only (**Fig. 4B,F)**. Post hoc analyses also revealed that changes in mean degree of IL_V1 and IL_MFG from baseline to the end of the follow-up were positively and negatively correlated to change of FMA-UE (*r* = 0.600, *p* = 0.002; *r* = -0.438, *p* = 0.037), respectively, indicating that the change of functional connectivity in both IL_V1 and in IL_MFG were associated with recovery of motor performance after the follow-up (**Fig. 4C,G**). Furthermore, in network space, for IL_V1, which is assigned to visual network, most of the degree differences (more connections to IL_V1 in the imVR group than in the Control group) are from the sensory/somatomotor hand, visual, auditory, cingulo-opercular task control, dorsal attention and ventral attention networks on the contralesional hemisphere (**Fig. 4D**). For IL_MFG, which is assigned to DMN, most of the degree differences (more connections to IL_MFG in the Control group than in the imVR group) are from the ventral attention, DMN, frontal-parietal task control, cingular-opercular task control networks on the contralesional hemisphere (**Fig. 4H**).

The results of the other 5 statistically significant regions found at the end of follow-up assessment are shown in **Supplementary Fig. 5** and detailed in the supplementary follow-up fMRI results as well.

## Discussion

In this study, we demonstrated the effectiveness of imVR-based UE rehabilitation in stroke patients in their subacute phase. Participants assigned to the imVR training showed statistically significant and clinically meaningful improvements in UE motor impairment and daily living activity compared with the conventional rehabilitation program up to at least 12 weeks post-intervention. Moreover, these improvements were associated with changes in brain functional connectivity in ipsilesional premotor cortex and ipsilesional dorsolateral prefrontal cortex immediately after the intervention, and in ipsilesional visual region and ipsilesional middle frontal gyrus at 12-weeks follow-up. In addition, we observed increases in motor recovery rates for the imVR group from baseline to post-intervention.

The clinical outcomes indicate that the imVR training has positive impacts on the recovery of UE function and activities of daily living (ADL) as assessed by the FMA-UE and BI, respectively. These beneficial effects may first be attributed to the imVR program with goal-orientated repetitive functional task practice, which encourages highly repetitive functional movements of the UE [46]. Repetition of task-specific movements is one of the fundamental principles of both motor learning and the production of cortical reorganization to improve motor function after a stroke [47-51]. Animal studies demonstrate that a high number of repetitions are necessary to induce behavioral changes after brain injury [52], and moreover, the amount of motor training correlates with motor recovery [53-55]. Similar evidence is also found in humans: positive associations between the amount of motor therapy and outcomes in meta-analyses of clinical trial data [56, 57], reduction of upper limb motor impairment with high-frequency physical therapy [58], and improvement of arm and hand function [59] and of functional UE in patients with chronic stroke [60] with a repetitive function task. imVR provides immediate motor feedback [61] and a strong sense of presence [62] during training, allowing for task-oriented repetitive exercises while changing the traditional treatment patterns which can feel boring for patients. These characteristics allow exercises to be repeated without causing fatigue and pain [63], leading to some potentially clinically important benefits compared with conventional rehabilitation and improvements in upper limb impairments that were translated into an improvement in ADL [3].

In addition, imVR may be beneficial for improving UE impairment. The imVR system used here incorporates a wide field of view, high resolution, head-mounted display (HMD), and auditory and tactile/force feedback [64] to provide an enriched environment (EE), exposing subjects to enhanced motor, sensory, cognitive, and social stimuli relative to a standard condition [65]. Animal studies suggest that exposure to EE promotes angiogenesis in ischemic brains [66], induces behavioral and neurobiological changes [67, 68], synaptic plasticity changes [69], and enhances sensorimotor function [65]. In addition, a clinical trial demonstrated that exposure to EE was associated with increased activity levels, improved function, mood and quality of life of stroke survivors in a cost-saving manner [70].

Improvements in UE performance from the imVR training evaluated after the intervention correlated with reorganization of the brain networks; i.e., more brain functional connectivity to the sensory/somatomotor hand network, particularly on the ipsilesional side. After the 3-week intervention, 2 of 4 regions that statistically significantly differed between the imVR and the Control groups, degree of IL-PMd, IL-M1, ipsilesional and assigned to the sensory/somatomotor hand network (**Table 3, Fig. 3A, Supplementary Fig. 4A**), had more connections to them from ventral and dorsal attention, frontal-parietal and cingulo-opercular task control, and visual networks. This implies that the unique features of imVR—for example, more repetitive functional movements encouraged, and more attention inspired—may have enhanced motor planning and learning, visual stimuli, and motor control. This phenomenon is more greatly manifested in the region of IL_PMd, where connections from the sensory/somatomotor hand network itself are also increased and the change in functional connectivity was associated with UE motor recovery (**Fig. 3C**) and ADL (**Supplementary Fig. 4E**), consistent with previous studies [71, 72]. The network reorganization was evidenced as well in the other 2 statistically significant regions, IL-, and CL-DLPFC, which belong to the DMN. In certain goal-oriented tasks, DMN was deactivated [73], negatively correlating to attention and control networks [74]. The imVR training might contribute to a deep engagement and immersion so that the brain functional connectivity to the DMN decreased. For IL- and CL-DLPFC, the networks to which the connections statistically significantly decreased are dorsal attention, fronto-parietal task control, and within network DMN, respectively. Thus, we postulate that the imVR training incites more functional connectivity to the ipsilesional motor network with less connectivity to DMN, in turn improving UE motor recovery.

After the 12-week follow-up, the effects of imVR training on brain network reorganization is wider-reaching, with 9 regions exhibiting statistically significant differences between the imVR and the Control group, among which 3 (IL_V1, CL_V1, and IL_LOC), 2 (IL_MFG and CL_mPFC), 3 (IL_PMv, CL_SPG, and IL_IFG), and 1 (CL_FP) regions were located in visual, DMN, dorsal attention, and fronto-parietal task control networks, respectively. The 3 regions in the visual network may play pivotal roles associated with improvements of UE motion recovery after follow-up, in that the connections to the regions were increased and the change of FC between the follow-up and the baseline in the IL_V1 was associated with UE motor recovery. We also observed that the networks to which the 3 regions connect are sensory/somatomotor hand, visual, auditory, cingulo-opercular task control, dorsal attention and ventral attention networks on the contralesional hemisphere. Studies suggested that V1 is involved in object recognition and representation, object localization, and vision-guided movement processing [75-77] and that LOC is functionally related to the hand area of primary somatosensory [78], which was shown in our previous study [79]. Compared with the Control group, imVR may create more functional connections from the visual network to the sensory/somaomotor hand and cingulo-opercular task control network on the contralesional side at least 5 months after stroke, potentially enhancing a compensatory mechanism for loss of neuro activities on the ipsilesional side. Another region that has increased functional connectivity is SPG on the contralesional side (CL_SPG). Previous studies have shown that SPG, a hub for the exchange of sensory and motor-related information and critical for guiding upper limb movements toward the target and adjusting the shape of the hand to grasp the target [80], involved in the control of body movement, visual movement, oculomotor nerve activity and the guidance of visual spatial attention [81-84], which is corroborated by our data in that the mean degree in CL_SPG is significantly increased from baseline to follow-up for the imVR group (Supplementary Fig. 5C). Additionally, for CL_SPG assigned to the dorsal attention network, most of the connectivity differences (more connections to CL_SPG in the imVR group than in the Control group) are from the sensory/somatomotor hand, visual, auditory and cingulo-opercular task control networks in both hemispheres (Supplementary Fig. 5E). Therefore, we conclude that the imVR training may strengthen the function of SPG, promoting the improvement of patients’ upper limb function after the imVR training guided patients to perform a large number of repeated upper limb flexion and extension and grasping movements, while concurrently giving visual feedback. A further mediation or path analysis of the SPG function will be done in the future study.

Our post-hoc analyses indicated that the brain reorganization of two groups changed over time. After 12 weeks post-intervention, as shown in **Supplementary Fig. 6**, except for IL_DLPFC, 3 of the 4 brain regions that statistically significantly differed in degree between the imVR and the Control groups at the post-invention, IL-PMd, IL-M1, and CL-DLPFC, were no longer statistically significant, instead reverting to the same level as the baseline, and near normal levels (see HCs in **Fig. 3B, Fig. 3F** and **Supplementary Fig. 4C** and **4D**), indicating that these three regions correlated with imVR rehabilitation intervention are specifically affected during the intervention. Additionally, none of the 9 regions that showed statistically significant degree differences after the 12-week follow-up maintained statistical significance at the post-intervention timepoint, as shown in **Supplementary Fig. 7**, implying that some effects of the imVR on specific brain reorganization were at a later stage of recovery. The statistically significant brain reorganization in the two post-stroke stages may be attributed to the rehabilitation intervention’s effect. The first weeks-to-months after a stroke is a critical time for neural plasticity [85, 86], within which most stroke survivors exhibit spontaneous recovery [87]. The degree difference between the imVR and the Control immediately after the intervention may primarily be caused by the rehabilitation intervention, which further affected brain at the follow-up period even though at this stage there is a higher proportion of spontaneous recovery, resulting in different spontaneous recovery phenotypes. Moreover, the different brain reorganization of the two groups at two stages of recovery may be associated with our observation, as shown in **Fig. 2**, that a significant increase of FMA-UE change rate for the imVR group after the training, but no statistically significant difference between post-intervention and follow-up, which needs to be elucidated in the future research.

With the previous discoveries that more brain functional connectivity was observed in the ipsilesional than in the contralesional side immediately post-intervention and vice versa after later follow-up, we conclude that the imVR training was able to not only increase the functional activity of the impaired hemisphere immediately after the training, but also assist UE recovery by reorganizing brain activity in the contralesional hemisphere in a later stroke recovery phase. While this brain reorganization in the two post-stroke stages seems complicated, our fMRI results have opened a new window to explore the most appropriate frequency, intensity, and type of imVR-based rehabilitation to promote motor recovery by observing and comparing changes in functional connectivity.

The first limitation of our study was that it lacked both masking of subjects for concealing possible placebo effect and a sham intervention to investigate whether the effects of the imVR rehabilitation are specific from the imVR training itself in that concealments of group allocation and intervention are rarely possible technologically for stroke rehabilitation [88]. The second limitation was its single-center design and limited sample size, as we could not divided the participants into groups according to the severity of motor dysfunction. Another limitation is the limitation of current immersive VR technology, which are unable to simulate all rehabilitation training. As a compromise, the subjects in the experimental group experienced a combination of 30-min normal rehabilitation and 30-min immersive VR rehabilitation.

In conclusion, this study has demonstrated that the imVR-based rehabilitation is a promising rehabilitation tool for improving the recovery of UE functional capabilities of subacute stroke patients, and that these improvements are associated with distinct brain reorganization at two post-stroke stages. The results of the study will be of benefit to future patients with stroke and may provide a new and improved method of stroke rehabilitation.

## Data Availability

The original contributions presented in the study are included in the article/supplementary material, further inquiries can be directed to the corresponding authors.

## Acknowledgements

We would like to thank Le Lin, Xinxin Lin, Huifang Li at the Department of Physical Medicine and Rehabilitation in the Second Affiliated Hospital of Wenzhou Medical University Management for helping with subject recruitment.

## Authors’ contributions

Songhe Jiang, Lejian Huang designed the study. Lejian Huang, Qianqian Huang, and Andrew D. Vigotsky analyzed the data and drafted the manuscript. Xixi Jiang, Pengpeng Gu, Wenzhan Tu, Linyu Fan, Bo Wu, Yun Jin and Qianqian Huang conducted the trial. All authors approve the final manuscript version to be published.

