## Supplementary for "Effectiveness of immersive VR-based rehabilitation on upper extremity recovery in subacute stroke: a randomized controlled trial"

**Table 1. Age, sex and motion comparison among imVRs, Controls, and HCs**

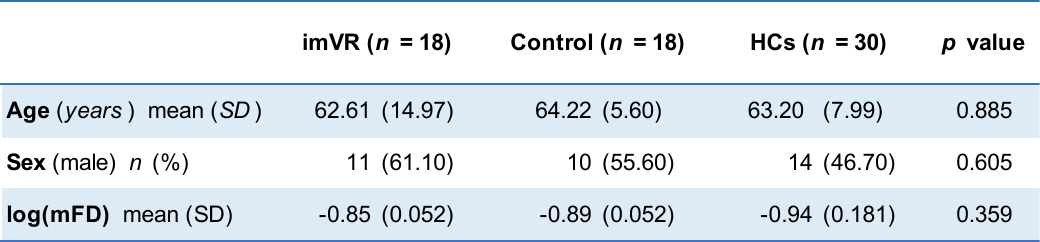

There was no significant difference between the imVR, Control and HCs groups for baseline characteristics. A One-way ANOVA test and Chi-square test were performed for continuous and categorical data, respectively. SD: Standard Deviation; imVR: immersive Virtual Reality; HCs: Healthy Controls; mFD: mean framewise displacement.

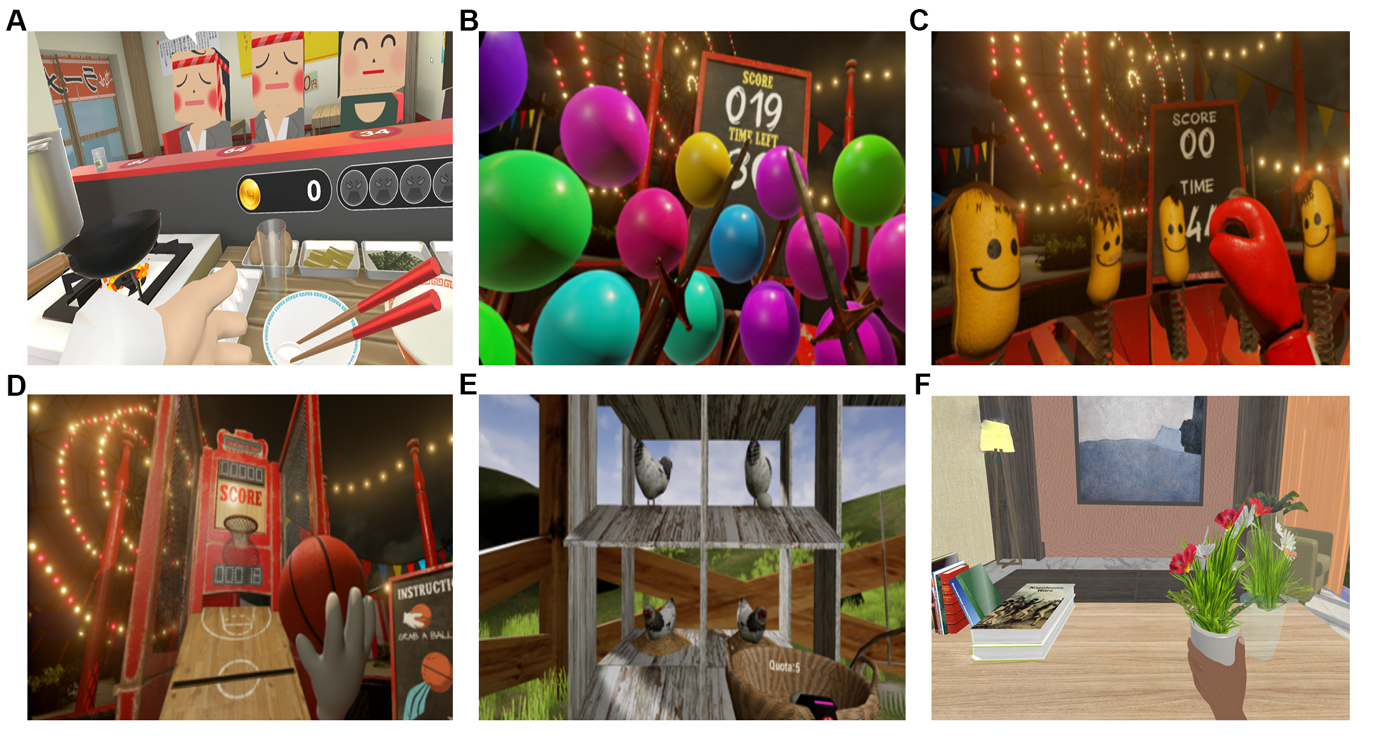

**Figure 1.** **Six imVR programs** (**A)** Frying dumplings and noodles by controlling a wok handle in a virtual kitchen. (**B)** Popping balloons by controlling a sword in a virtual fencing hall. (**C)** Punching with dolls by controlling a big fist in a virtual boxing arena. (**D)** Playing basketball in a virtual court, in which a controller shoots the ball, and the height and distance is varied over time. (**E)** Collecting eggs into a virtual basket by a controller. (**F)** Tidying up a desk and moving objects to a designated position in a virtual office.

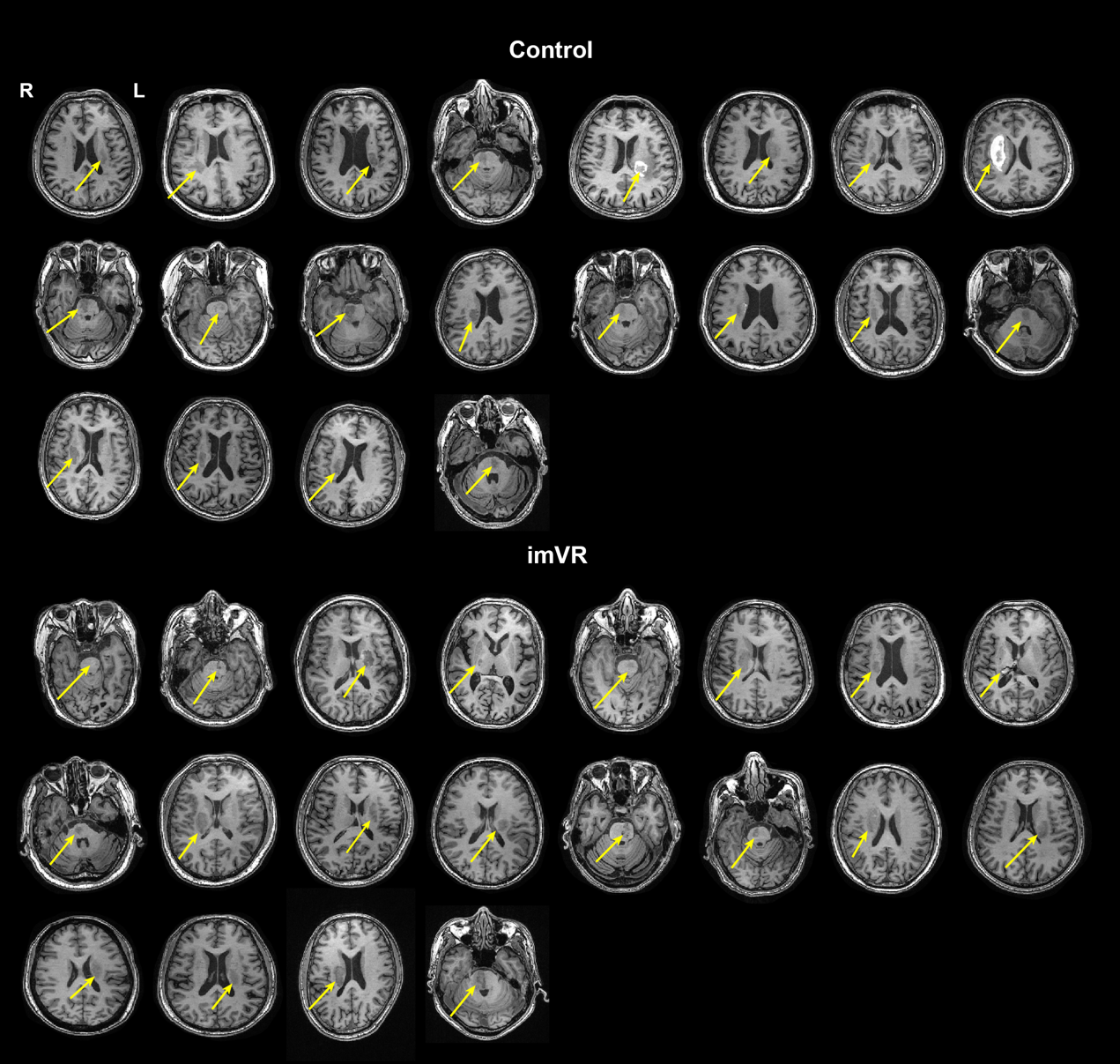

**Figure 2.** Yellow arrows indicate the largest lesion area of each stroke patient at the axial cross section for the imVR (bottom) and the Control (top) groups. R and L represent right and left hemisphere of brain, respectively.

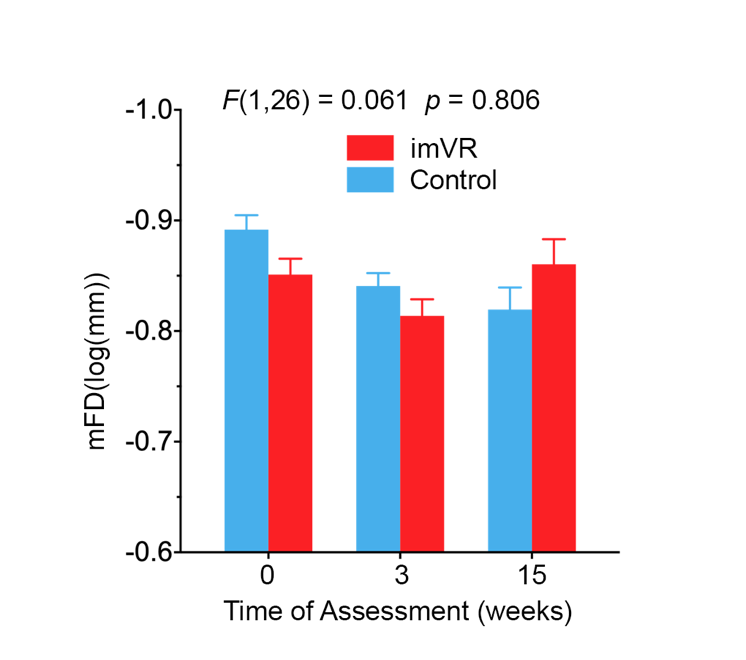

**Figure 3.** Bar graph (mean and SE) shows no significant difference of brain motion during scanning, represented by *log*(mFD), across three assessments between the imVR and the Control groups. mFD: mean frame-wise displacement.

**Supplementary post-intervention fMRI results**

After cluster-correction (*t* > 3.5 and *p* < 0.01), the imVR group exhibited higher degree in IL_PMd (*p* = 0.008) and IL_M1 (*p* = 0.003) regions (**Fig. 4A**), and lower degree in IL_and CL_DLPFC (*p* = 0.003; *p* < 0.001) regions at the end of intervention compared to the Control group (Week 3) (**Fig.** **4B**). Post hoc analysis indicates that the imVR group had greater mean degree in IL_M1 (*t*_28_ = 4.46, *p* < 0.001) and lower degree in CL_DLPFC (*t*_28_ = -4.28, *p* < 0.001) than the Control group at the end of the intervention (**Fig.** **4C, D**). Compared with HCs, the mean degree in IL_M1 was decreased (*p* = 0.037) from baseline to post-intervention for the Control group, while the mean degree in CL_DLPFC is significantly increased (*p* = 0.015) in the Control group. Post hoc analyses also revealed that the change of mean degree in IL_PMd between post-intervention and baseline was positively correlated to the change of BI (*r* = 0.575, *p* = 0.004), indicating that the change of functional connectivity in IL_PMd was associated with recovery of activities of daily living (ADL) after the intervention (**Fig. 4E**). Furthermore, in network space, for IL_M1, which is assigned to the sensory/somatomotor hand network, most of the degree difference (more connections to IL_M1 in the imVR group than in the Control group) are from sensory/somatomotor hand, ventral attention, dorsal attention, DMN, fronto-parietal task control and cingulo-opercular task control networks on the ipsilesional hemisphere (**Fig. 4F, G**). For CL_DLPFC, which is assigned to the DMN, most of the differences (more connections to CL_DLPFC in the Control group than in the imVR group) were from ventral attention, fronto-parietal task control and DMN on the contralesional hemisphere, and DMN, cingulo-opercular task control and frontal-parietal task control network on ipsilesional hemisphere.

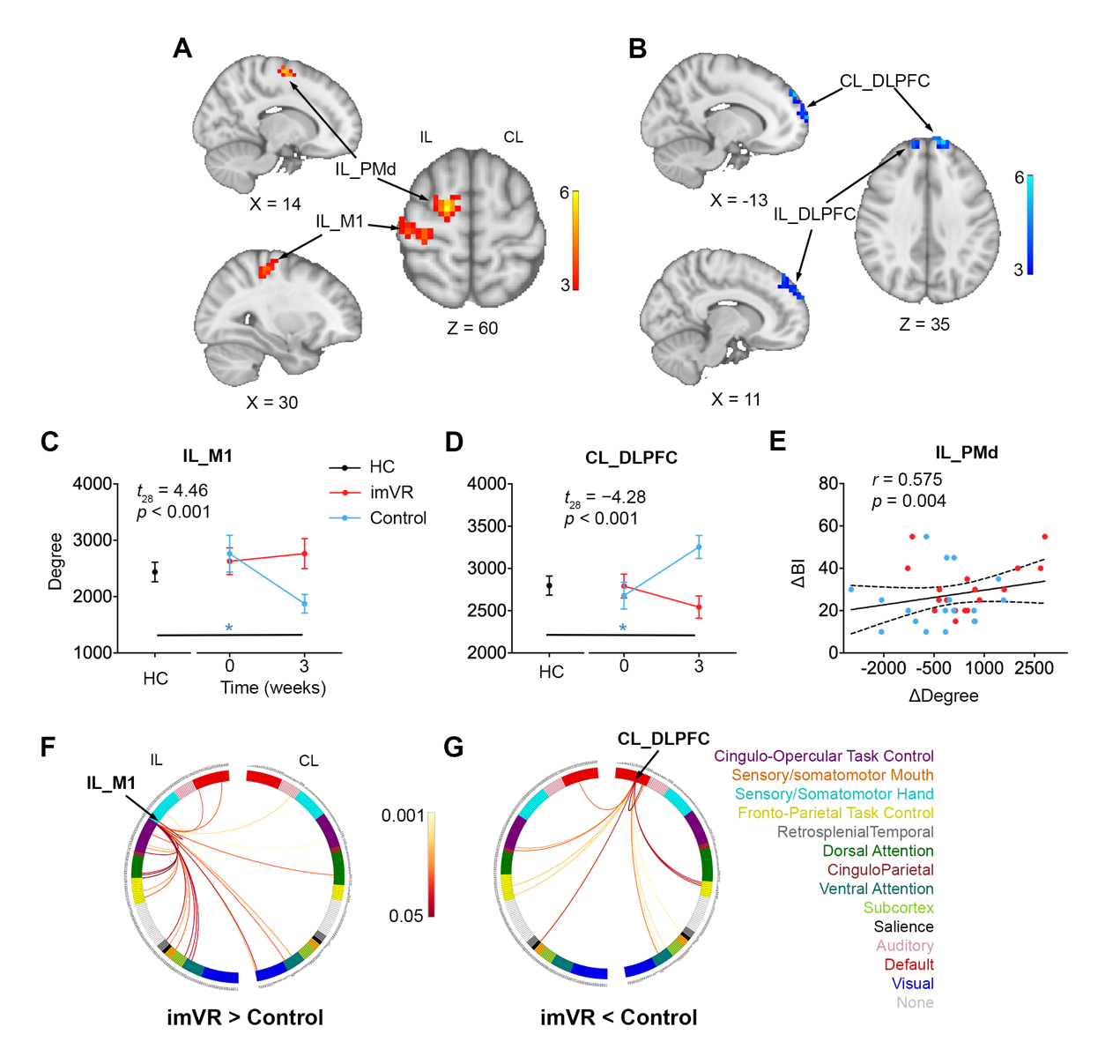

**Figure 4. Brain properties at the end of the intervention.** **(A)** imVR had greater degree in IL_PMd and IL_M1 compared with the Control group (cluster-corrected, *t* > 3.5, *p* < 0.01). **(B)** imVR had lower degree in CL_ DLPFC and IL_DLPFC regions compared with the Control group (cluster-corrected, *t* > 3.5, *p* < 0.01). **(C)** Post hoc analysis revealed that imVR had greater mean degree in IL_M1 than the Control group (*t*_28_ = 4.46, *p* < 0.001) at the end of the intervention. Compared with HCs, the Control group’s mean degree in IL_M1 decreased (*p* = 0.037) from baseline to post-intervention. Data plotted as mean ± SE. **(D)** Post hoc analysis also revealed that imVR had lower mean degree in CL_DLPFC than the Control group (*t*_28_ = -4.28, *p* < 0.001) at the end of the intervention. Compared with HCs, the mean degree in CL_DLPFC increased (*p* = 0.015) from baseline to post-intervention for the Control group. **(E)** IL_PMd degree changes positively correlated with changes in BI (*r* = 0.575, *p* = 0.004) from baseline to the end of the intervention. **(F)** The circular plot shows the difference in functional connections to IL_M1 between the imVR and Control groups (*p* < 0.05) in the network space. The IL_M1 region assigned to the sensory/somatomotor hand network. Most of the differences in connections are from the fronto-parietal task control, ventral attention, dorsal attention, default and cingulo-opercular task control networks on the ipsilesional hemisphere. **(G)** The circular plot shows the difference in functional connections to CL_LDPFC between the imVR and Control groups (*p* < 0.05) in the network space. The CL_LDPFC region is assigned to the DMN. Most of the differences in connections are from ventral attention, fronto-parietal task control and DMN on the contralesional hemisphere, and DMN, cingulo-opercular task control and frontal-parietal task control network on ipsilesional hemisphere.

ΔDegree = post-intervention degree minus baseline degree; ΔBI = post-intervention BI minus baseline BI; PMd: Dorsal Premotor Cortex; M1: Primary Motor Cortex; DLPFC: Dorsolateral Prefrontal Cortex; IL: Ipsilesional; CL: Contralesional.

**Supplementary follow-up fMRI results**

At the end of the follow-up (Week 15), after cluster-correction (*t* > 3.5 and *p* < 0.01), compared with the Control group, the imVR group exhibited higher degree in IL_V1 (*p* = 0.002), CL_V1 (*p* < 0.001), CL_SPG (*p* < 0.001) and IL_LOC (*p* < 0.001) (**Fig. 5A**), and lower degree in IL_MFG (*p* < 0.001), IL_PMv (*p* = 0.004), IL_IFG (*p* < 0.001), CL_mPFC (*p* < 0.001) and CL_FP (*p* < 0.001) regions at the end of follow-up (Week 15) (**Fig. 5B**).

As shown in **Fig. 5C** and **Fig. 5D**, post hoc analysis indicates the imVR group presents higher mean degree in CL_V1 (*t*_20_ = 2.87, *p* = 0.01), CL_SPG (*t*_20_ = 4.65, *p* < 0.001) and IL_LOC (*t*_20_ = 5.06, *p* < 0.001) and lower degree in IL_MFG (*p* < 0.001), IL_PMv (*p* = 0.004), IL_IFG (*p* < 0.001), CL_mPFC (*p* < 0.001) and CL_FP (*p* < 0.001) regions than the Control group at the end of the follow-up. Compared with HCs, the mean degree in CL_V1 and IL_LOC is decreased (*p* < 0.001), and the mean degree in CL_SPG is increased (*p* = 0.001) from baseline to follow-up for the Control and imVR groups, respectively. Compared with HCs, the mean degree in IL_PMv, IL_IFG and CL_mPFC decreased (*p* < 0.001, *p* < 0.001, *p* < 0.001) from baseline to follow-up for the imVR group, while the mean degree in IL_PMv and CL_FP increased (*p* = 0.034, *p* = 0.001) for the Control group. Furthermore, in network space, as shown in **Fig. 5E,** for CL_V1 and IL_LOC regions, which are assigned to the visual network, and CL_SPG assigned to the dorsal attention network, most of the degree differences (more connections to CL_V1, CL_SPG and IL_LOC in the imVR group than in the Control group) are from the sensory/somatomotor hand, visual, auditory, cingulo-opercular task control, dorsal attention and ventral attention networks on the contralesional hemisphere. The IL_PMv and IL_IFG regions assigned to the dorsal attention network and most of the differences (more connections to IL_PMv and IL_IFG in the Control group than in the imVR group) are from the default, dorsal attention, cingular-opercular task and sensory/somatomotor hand networks on the contralesional hemisphere. CL_mPFC assigned to the default network and most of the differences (more connections to CL_mPFC in the Control group than in the imVR group) are from the ventral attention, default, fronto-parietal task control, cingular-opercular task control networks on the ipsilesional hemisphere. CL_FP assigned to the fronto-parietal task control network and most of the differences (more connections to CL_FP in the Control group than in the imVR group) are from the dorsal attention, default, fronto-parietal task control, cingular-opercular task control networks on the ipsilesional hemisphere.

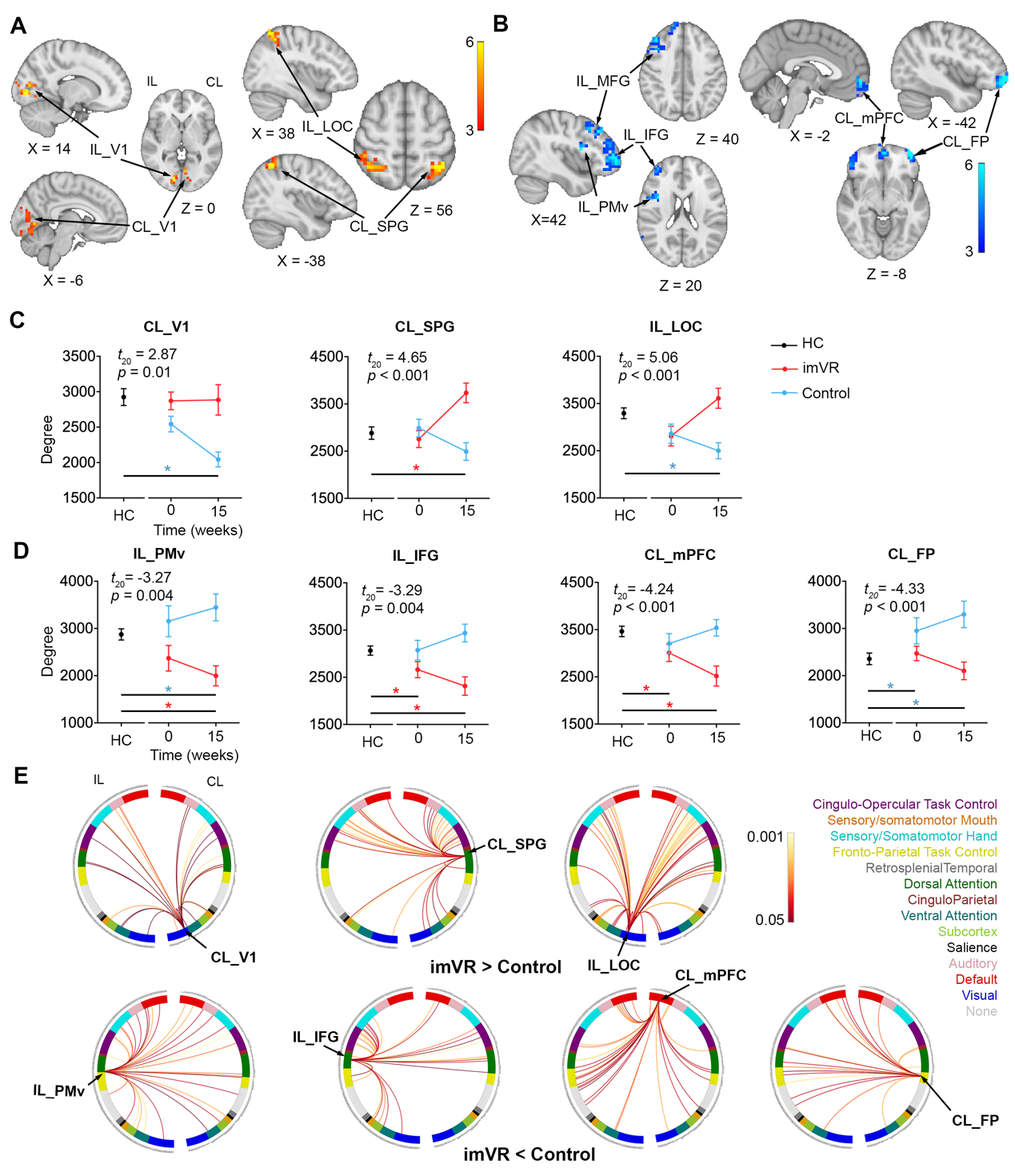

**Figure 5. Brain properties at the end of follow-up**. **(A)** imVR group had greater degree in IL_V1, CL_V1, CL_SPG and IL_LOC regions at the end of the follow-up compared with the Control group (cluster-corrected, *t* > 3.5, *p* < 0.01). **(B)** imVR group had lower degree in IL_MFG, IL_PMv, IL_ IFG, CL_mPFC and CL_FP regions at the end of intervention compared with the Control group (cluster-corrected, *t* > 3.5, *p* < 0.01). **(C)** Post hoc analysis indicates imVR had greater mean degree in CL_V1 (*t*_20_ = 2.87, *p* = 0.01), CL_SPG (*t*_20_ = 4.65, *p* < 0.001) and IL_LOC (*t*_20_ = 5.06, *p* < 0.001) than the Control group at the end of the follow-up. Compared with HCs, the mean degree in CL_V1 and IL_LOC decreased (*p* < 0.001) from baseline to the post-intervention for the Control group, while the mean degree in IL_SPG increased (*p* = 0.001) from baseline to the end of follow-up for the imVR group. Data plotted as mean ± SE. **(D)** Post hoc analysis indicates imVR had lower mean degree in IL_PMv (*t*_20_ = -3.27, *p* = 0.004), IL_IFG (*t*_20_ = -3.29, *p* = 0.004), CL_mPFC (*t*_20_ = -4.24, *p* < 0.001 ) and CL_FP (*t*_20_ = -4.33, *p* < 0.001) than the Control group at the end of the follow-up. Compared with HCs, the mean degree in IL_PMv (*p* < 0.001), IL_IFG (*p* < 0.001) and CL_mPFC (*p* < 0.001) decreased from baseline to follow-up for the imVR group, while the mean degree in IL_PMv (*p* < 0.001) and CL_FP (*p* < 0.001) is increased from baseline to follow-up for the Control group. **(E)** Functional connections to CL_V1, CL_SPG and IL_LOC differed between the Control and imVR groups (*p* < 0.05) in the network space. CL_V1 and IL_LOC regions were assigned to the visual network and CL_SPG assigned to the dorsal attention network. Most of the differences of connections are from the sensory/somatomotor hand, visual, auditory, cingulo-opercular task control, dorsal attention and ventral attention networks on the contralesional hemisphere. Circular plot shows the difference in functional connections to IL_PMv, IL_IFG, CL_mPFC and CL_FP, which differed between the Control and imVR groups (*p* < 0.05) in the network space. IL_PMv and IL_IFG assigned to the dorsal attention network and most of the differences in connections are from the default, dorsal attention, cingular-opercular task and control sensory/somatomotor hand networks on the contralesional hemisphere. CL_mPFC and CL_FP were assigned to the default and fronto-parietal task control network, respectively. Most of the differences in connections in both CL_mPFC and CL_FP are from the dorsal attention, default, fronto-parietal task control, cingular-opercular task control networks on the ipsilesional hemisphere.

V1: Primary Visual Cortex; SPG: Superior Parietal Gyrus; LOC: Lateral Occipital Cortex; MFG: Middle Frontal Gyrus; PMv: ventral premotor cortex; IFG: Inferior frontal gyrus; mPFC: medial Prefrontal Cortex; FP: Frontal Pole; IL: Ipsilesional; CL: Contralesional.

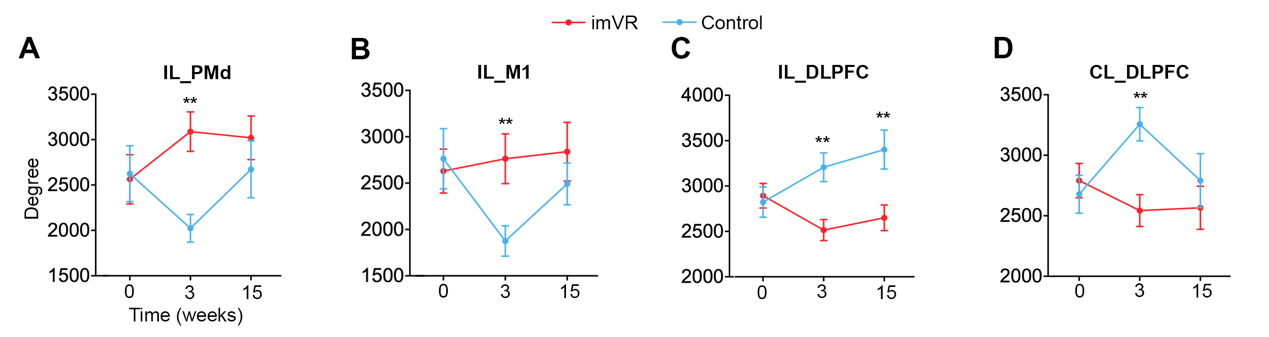

**Figure 6.** **Time-dependent changes in degree extracted from the 4 significant regions found at the post-intervention timepoint.** Degree was compared between imVR and Control groups, at week 0, week 3, week 15. Data plotted as mean ± SE. PMd: Dorsal Premotor Cortex; M1: Primary Motor Cortex; DLPFC: Dorsolateral Prefrontal Cortex; IL: Ipsilesional; CL: Contralesional;imVR: immersive virtual reality.

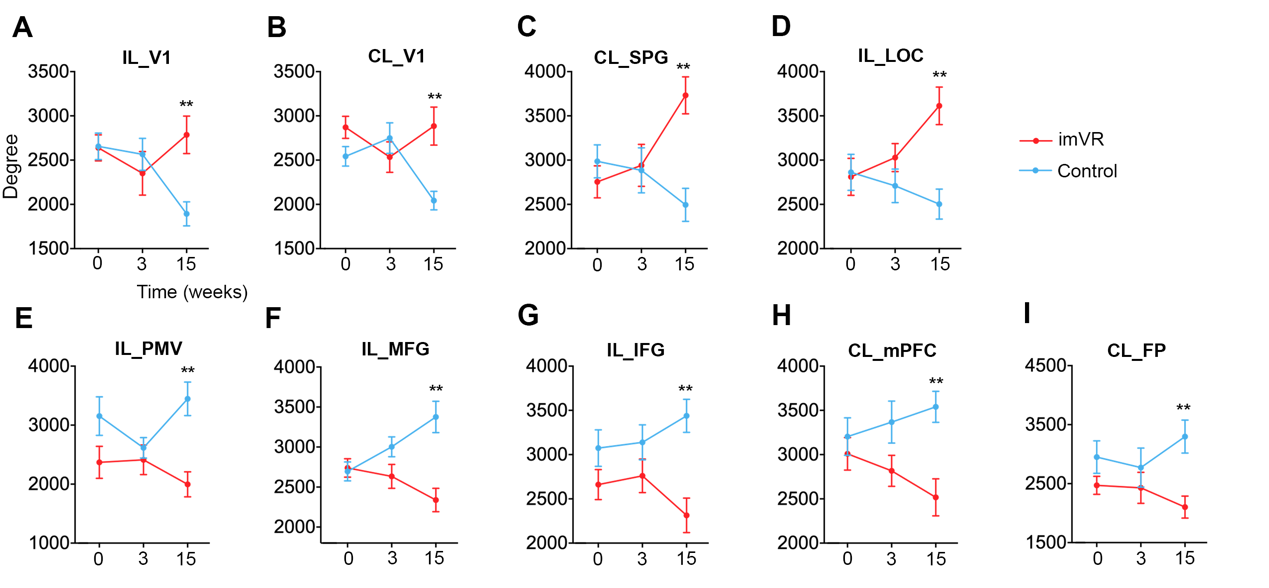

**Figure 7.** **Time-dependent changes in degree for the 9 significant regions found at follow-up.**  V1: Primary Visual Cortex; SPG: Superior Parietal Gyrus; LOC: Lateral Occipital Cortex; MFG: Middle Frontal Gyrus; PMv: ventral premotor cortex; IFG: Inferior frontal gyrus; mPFC: medial Prefrontal Cortex; FP: Frontal Pole; CL: Contralesional; IL: Ipsilesional; imVR: immersive virtual reality.
